# Effect of Odor Stimulations on Physical Activity: A Systematic Review

**DOI:** 10.1101/2022.10.04.22280694

**Authors:** Mathieu Cournoyer, Alice Maldera, Alexandre-Charles Gauthier, Fabien Dal Maso, Marie-Eve Mathieu

**Affiliations:** École de Kinésiologie et des Sciences de l’Activité Physique de la Faculté de Médecine, Université de Montréal, 2100 Edouard Montpetit Blvd #8223, Montreal, QC H3T 1J4, Canada; Centre Interdisciplinaire de Recherche sur le Cerveau et l’Apprentissage, Montréal, QC, Canada; Centre de Recherche du CHU Sainte-Justine, 2100 Edouard Montpetit Blvd #8223, Montreal, QC H3T 1J4, Canada

**Keywords:** exercise, odors, physical activity, ergogenic effect, performance, smell

## Abstract

Fewer and fewer people are reaching physical activity recommendations. Therefore, it seems important to make the practice of physical activity more enjoyable to increase the participation rate. Several environmental factors have been studied to see their impact on sports practice, and some studies investigated the effect of odors. This systematic review aims to provide a thorough view of the literature on the effect of different odors on physical activity. The search strategy consisted of using index terms and keywords in MEDLINE, EMBASE, EBM Reviews – Cochrane Central Register of Controlled Trials, CINAHL, SPORTDiscus, and Web of Science search engine. Data from 19 studies that included 421 participants revealed that the odors had different results on strength, cardiovascular, precision, and postural balance tasks depending on the odors’ exposition. Among results, an important distinction was made between pleasant and unpleasant odors. Therefore, pleasant odors had better results on physical activity by improving participants’ feeling. Even though this review clarified evidence about the effect of odors on physical activity, better methodological consistency is needed across studies such as the odor administration method to produce more meaningful results.

## 1 Introduction

Physical activity (PA), as described by the World Health Organization, represents “any bodily movement produced by skeletal muscles that requires energy expenditure” [1]. This type of therapeutic intervention has proven its efficacy in preventing cardiovascular, metabolic, and neurologic diseases [1]. In the past decades, the number of people who have not reached PA recommendations, generally established at a minimum of 150 minutes of aerobic exercise at a moderate to high intensity per week [1], has consistently increased. Statistics Canada showed that in 2021, almost half of the Canadian population over 12 years old self-declared that they did not reach those recommendations [2]. This observation shows the importance of finding strategies to make PA more appealing by promoting pleasant sensations during its practice.

Thereby, many factors can affect the practice of PA, such as external stimuli, which can change the appreciation of the experiences and dictate our feelings on a daily basis [3]. For example, some odors (stimuli) might positively affect our pain perception by reducing its intensity [3, 4]. Specifically, pleasant odors may have the ability to reduce sympathetic nervous system activity, which in turn could reduce overall pain feeling. However, it is important to note that an unpleasant odor would have the opposite effect and could further activate the nervous system [3, 5]. Another study exposed the ergogenic effect of mint and cinnamon by improving the arousal level when driving a car, meaning participants were more focused on the task [3, 6]. It is thus possible to question whether similar findings can be sought by athletes to improve their sports performance.

A deeper understanding of the effects of certain odors on PA would be beneficial to highlight the mechanisms modulating healthy lifestyle habits. This systematic review, therefore, aims to highlight the scientific consensus on the influence of certain odors on the mechanisms related to the practice of PA. We will explore the effect of some odors such as peppermint, lavender, and citrus on various types of PA, such as strength, cardiovascular, precision, and balance tasks.

## 2 Methods

### 2.1 Search Strategy

This systematic review followed the “Preferred Reporting Items for Systematic Reviews and Meta-Analysis” (PRISMA) guidelines [7]. Six databases were used to perform the search: Medline (1946-present), Embase (1974-present), EBM Reviews – Cochrane Central Register of Controlled Trials (1991-present), CINAHL Plus with Full Text (1937-present), SportDiscus with Full Text (1930-present) and Web of Science (1945-present). The last research was performed on December 22, 2021. The following keywords were used to search the concept of: “aerobic training” OR exercise* OR “physical activity” OR “physical training” OR “resistance training” OR sport* OR “strength training” OR “weight training” OR weightlifting OR “weight lifting” OR running OR football OR hockey OR athletes OR exertion OR “athletic performance”. For olfaction, the following keywords were used: odor OR odour OR aroma OR fragrance* OR smelling OR smell OR olfactory OR olfaction OR normosmic OR normosmia OR hyposmic OR hyposmia OR anosmic OR anosmia OR odorant* OR sniff OR sniffing.

Included studies had to meet the following inclusion criteria: (1) investigate the effect of certain odors on PA; (2) involve humans, both adults and children; (3) essential oils or odors were administered nasally. On the other hand, studies were not included if (1) olfactory acuity had changed following head trauma or surgery; (2) studies were not completed or published; (3) studies were written in a language other than English or French.

### 2.2 Study Selection

Figure 1 shows the evolution of the number of articles retained and excluded at each stage. Duplicates were removed after studies identification. The first screening of articles was done only using titles by two authors (MC, AD). The second screening of articles was done using abstracts by two authors (MC, AD). Subsequently, a final sorting was done by two authors (MC, AD) using the full texts of the remaining studies. All authors’ conflicts were discussed internally and resolved by a third author (ACG).

**Figure 1.**
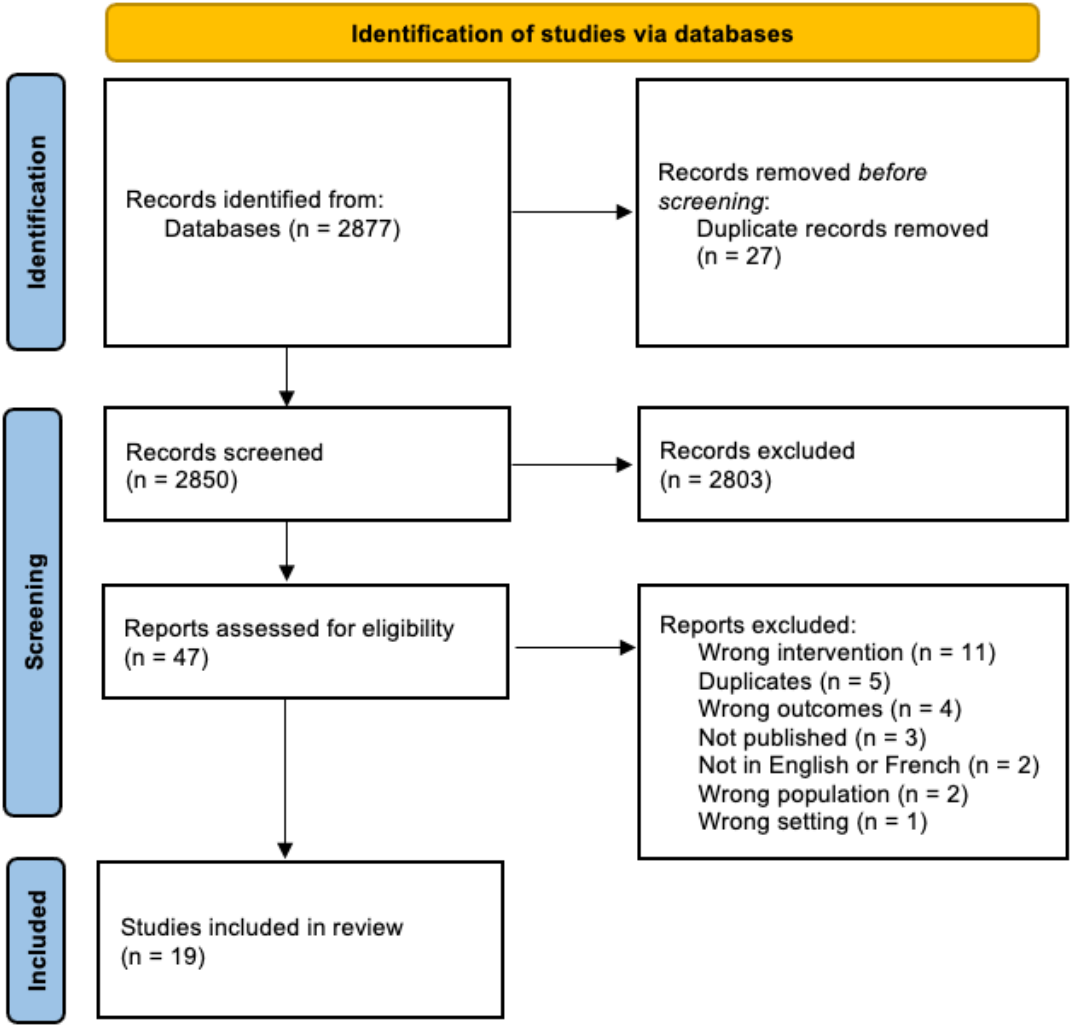
Flowchart of studies selection from their identification to final inclusion

### 2.3 Data Extraction and Analyses

Articles included following the selection process were read and analyzed by two authors (MC, AD). The data were processed by MC, extracted by MC and validated independently by AD. If differences were present, the authors reached a consensus following a discussion. If necessary, a third author (ACG) was consulted to make the decision. Data from included studies are summarized in Table 1, and their protocols are summarized in Table 2.

**Table 1.**
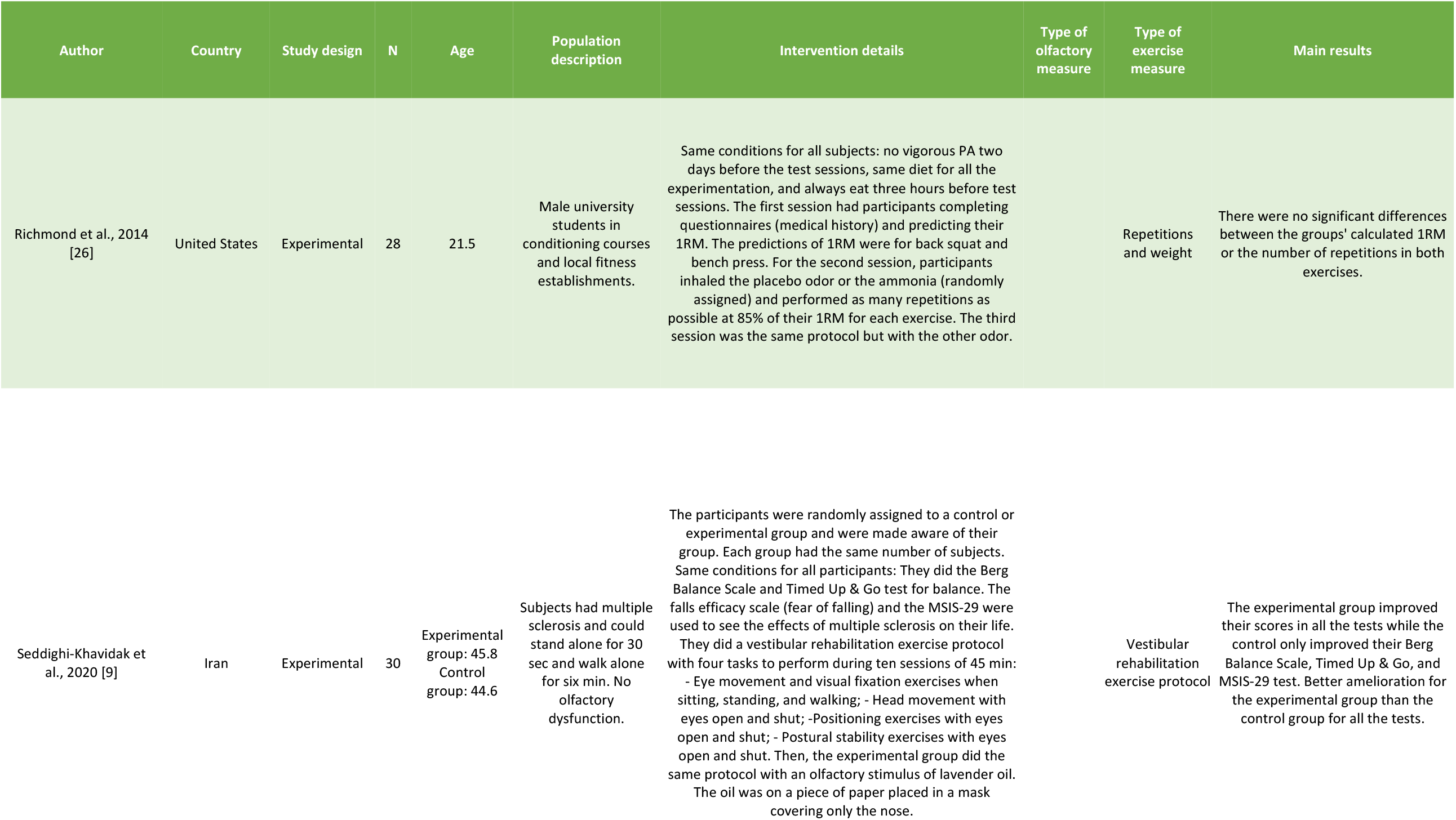
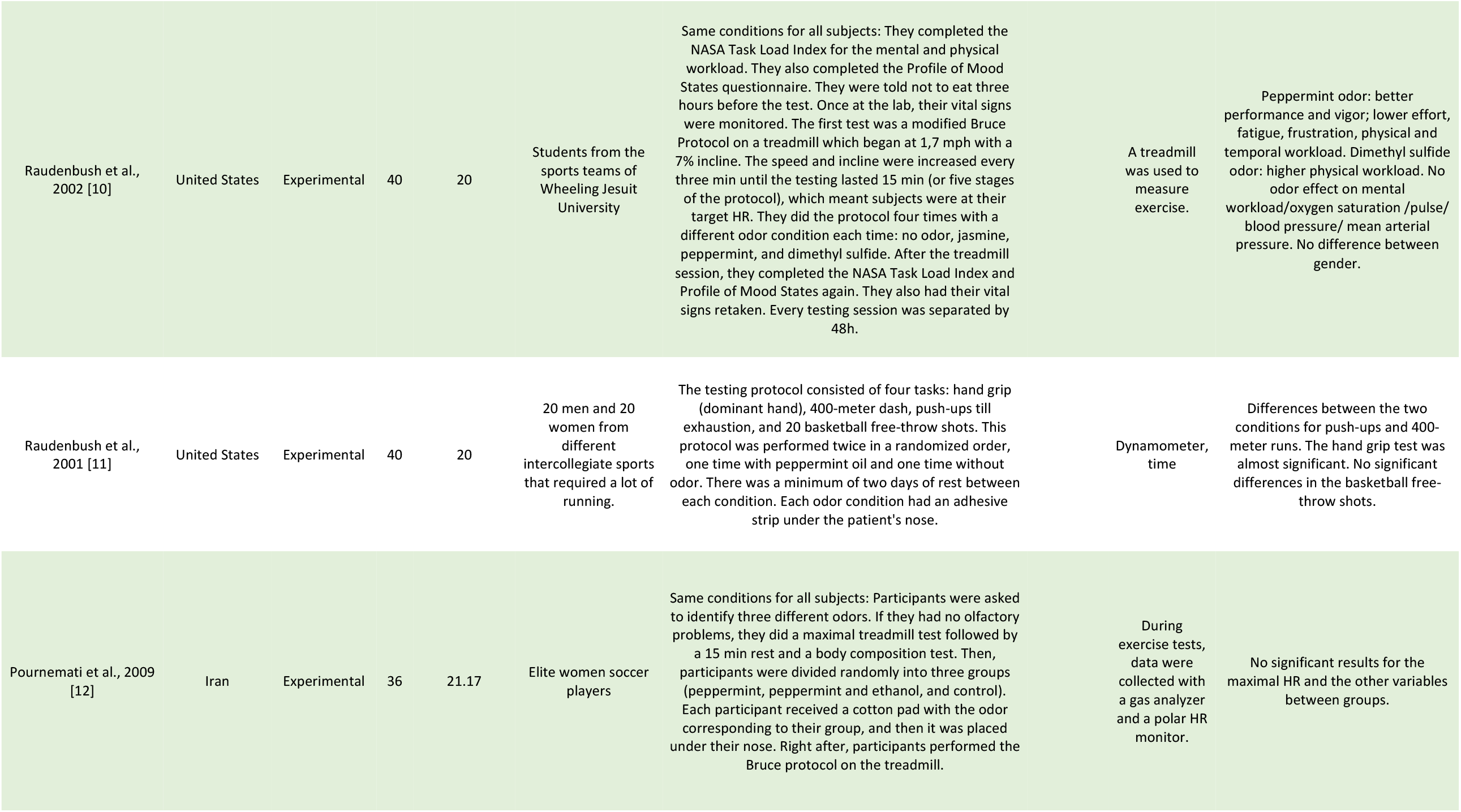
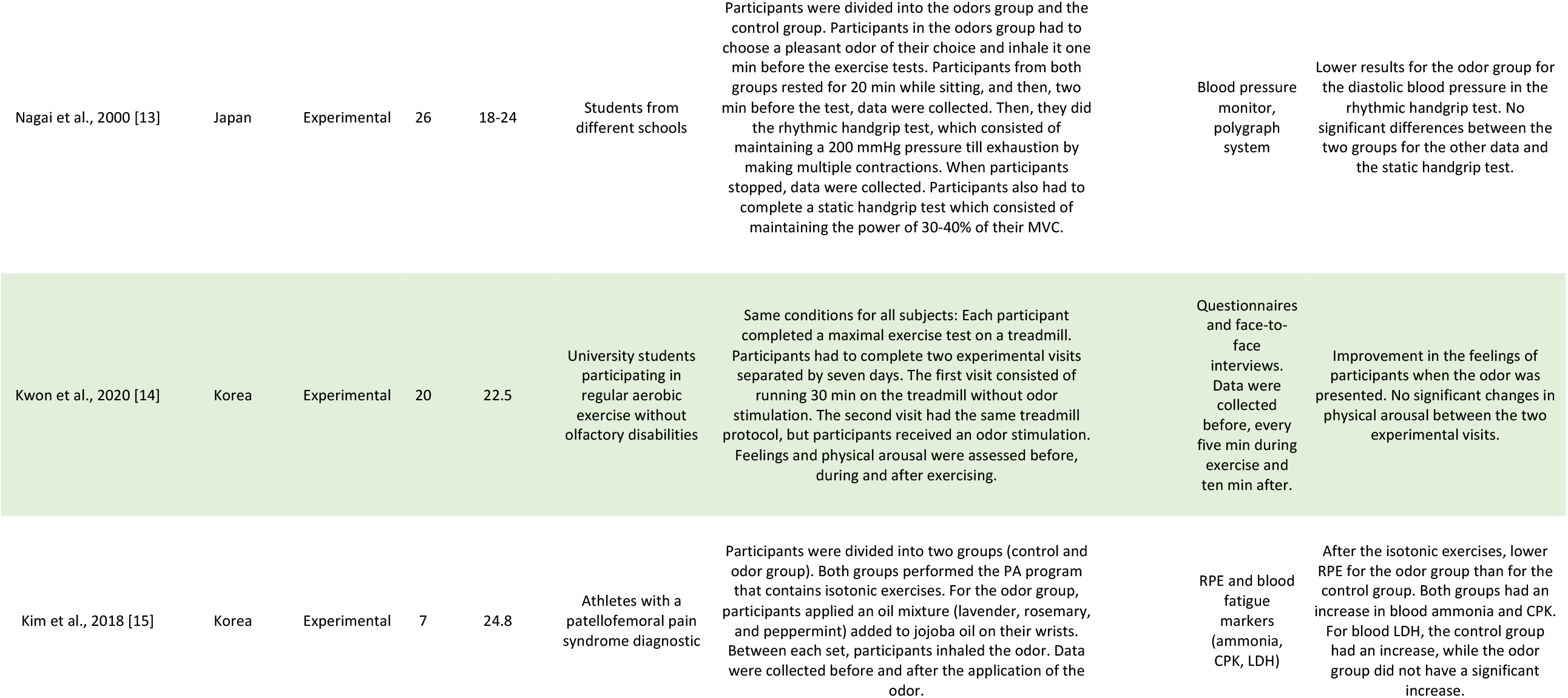
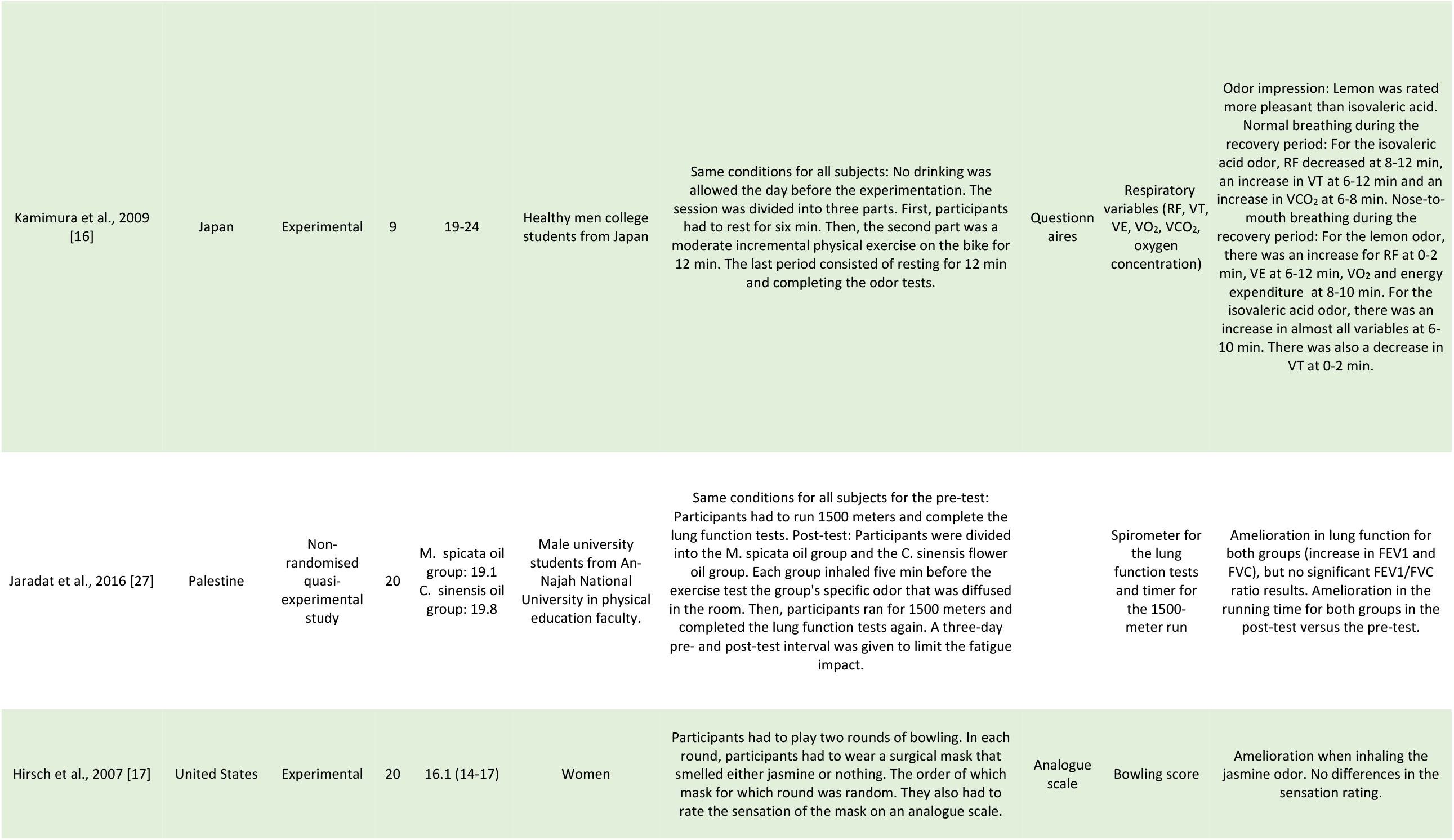
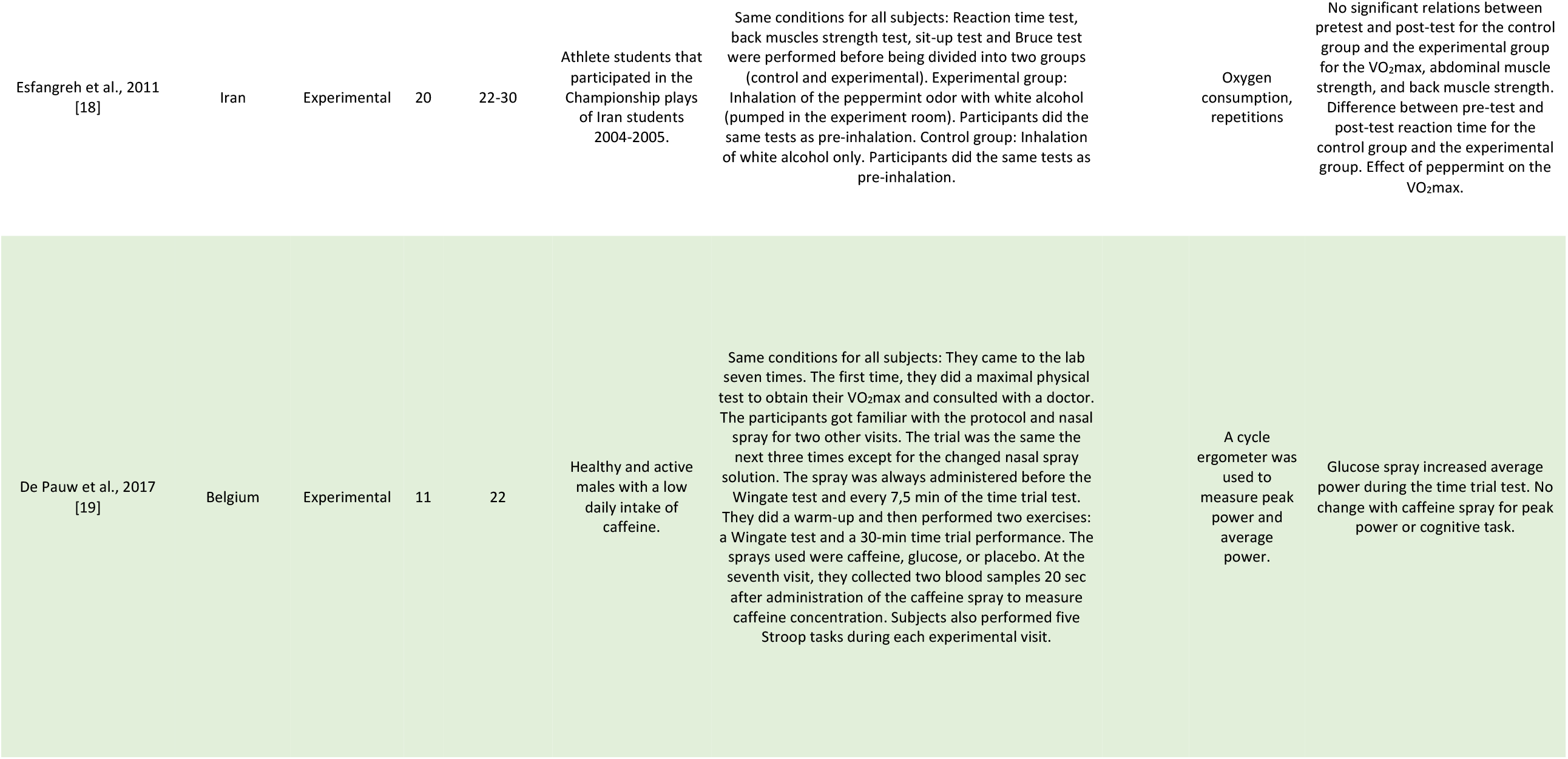
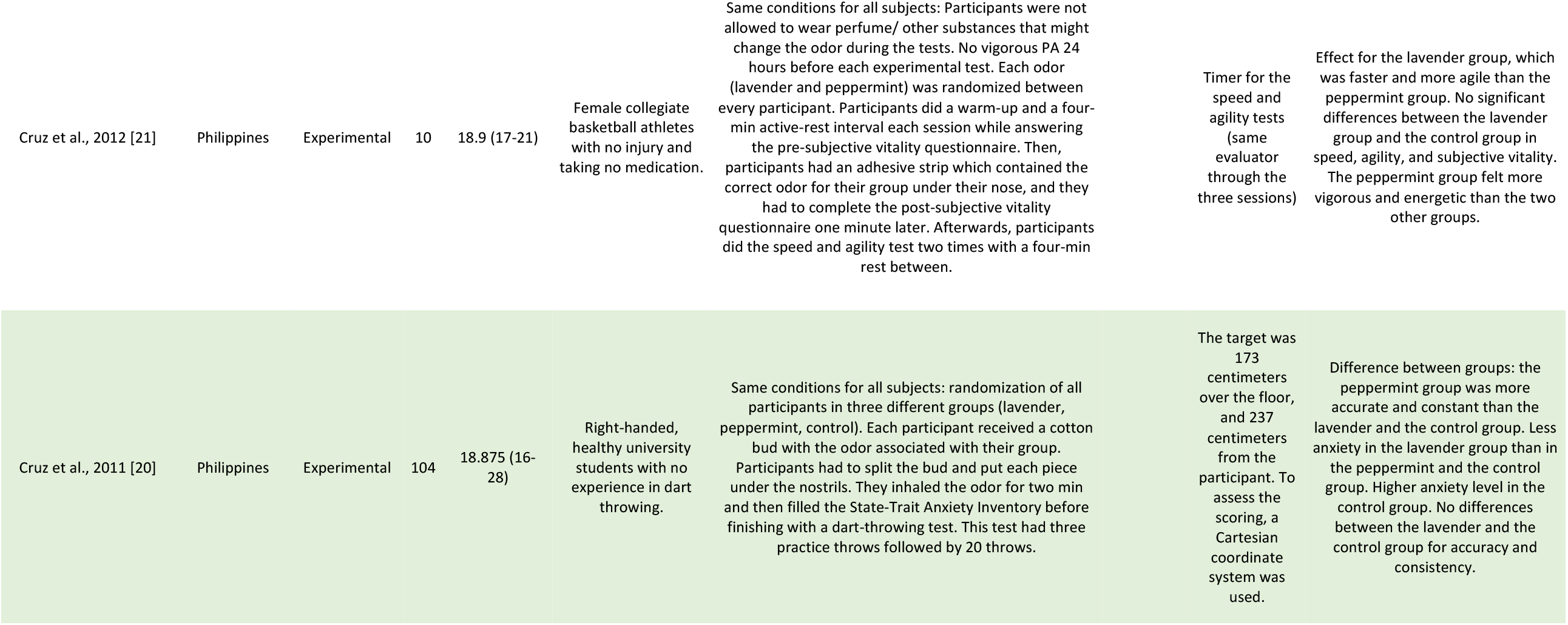
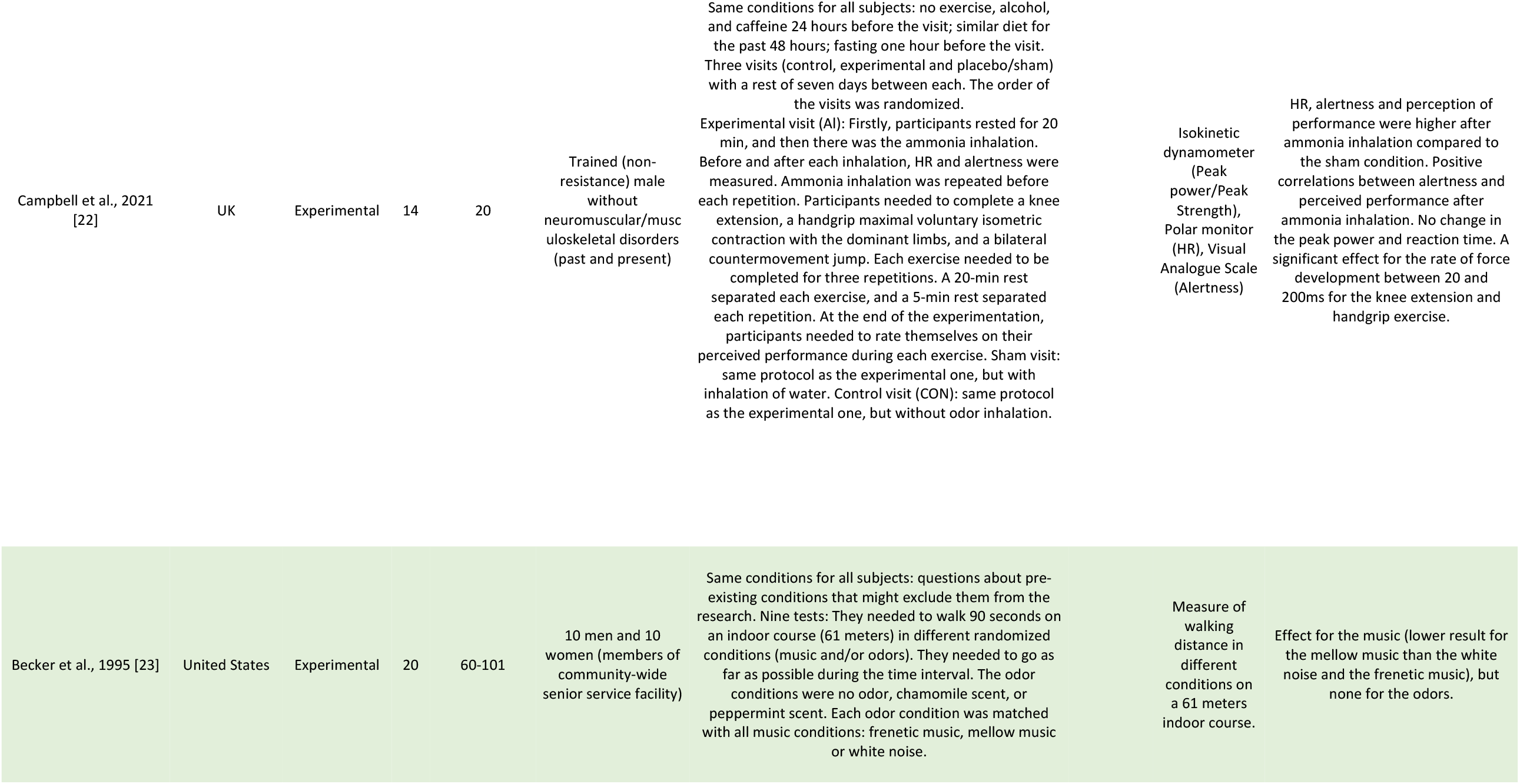
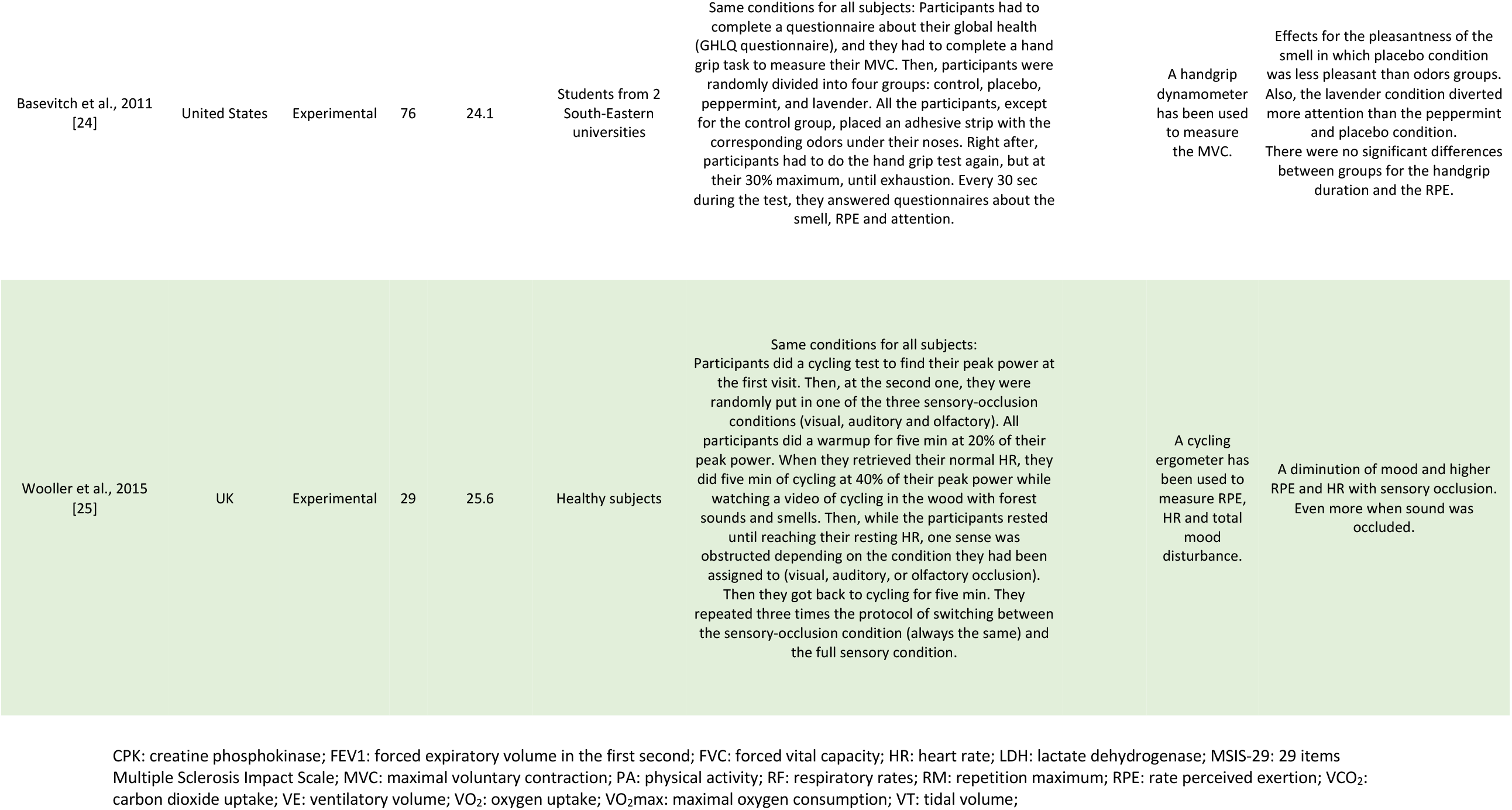
Included studies’ characteristics

| Author | Country | Study design | N | Age | Population description | Intervention details | Type of olfactory measure | Type of exercise measure | Main results |
| --- | --- | --- | --- | --- | --- | --- | --- | --- | --- |
| Richmond et al., 2014 [26] | United States | Experimental | 28 | 21.5 | Male university students in conditioning courses and local fitness establishments. | Same conditions for all subjects: no vigorous PA two days before the test sessions, same diet for all the experimentation, and always eat three hours before test sessions. The first session had participants completing questionnaires (medical history) and predicting their 1RM. The predictions of 1RM were for back squat and bench press. For the second session, participants inhaled the placebo odor or the ammonia (randomly assigned) and performed as many repetitions as possible at 85% of their 1RM for each exercise. The third session was the same protocol but with the other odor. |  | Repetitions and weight | There were no significant differences between the groups' calculated 1RM or the number of repetitions in both exercises. |
| Seddighi-Khavidak et al., 2020 [9] | Iran | Experimental | 30 | Experimental group: 45.8<br>Control group: 44.6 | Subjects had multiple sclerosis and could stand alone for 30 sec and walk alone for six min. No olfactory dysfunction. | The participants were randomly assigned to a control or experimental group and were made aware of their group. Each group had the same number of subjects. Same conditions for all participants: They did the Berg Balance Scale and Timed Up & Go test for balance. The falls efficacy scale (fear of falling) and the MSIS-29 were used to see the effects of multiple sclerosis on their life. They did a vestibular rehabilitation exercise protocol with four tasks to perform during ten sessions of 45 min:<br>- Eye movement and visual fixation exercises when sitting, standing, and walking;<br>- Head movement with eyes open and shut;<br>- Positioning exercises with eyes open and shut;<br>- Postural stability exercises with eyes open and shut. Then, the experimental group did the same protocol with an olfactory stimulus of lavender oil. The oil was on a piece of paper placed in a mask covering only the nose. |  | Vestibular rehabilitation exercise protocol | The experimental group improved their scores in all the tests while the control only improved their Berg Balance Scale, Timed Up & Go, and MSIS-29 test. Better amelioration for the experimental group than the control group for all the tests. |

|  |  |  |  |  |  |  |  |  |
| --- | --- | --- | --- | --- | --- | --- | --- | --- |
| Raudenbush et al., 2002 [10] | United States | Experimental | 40 | 20 | Students from the sports teams of Wheeling Jesuit University | Same conditions for all subjects: They completed the NASA Task Load Index for the mental and physical workload. They also completed the Profile of Mood States questionnaire. They were told not to eat three hours before the test. Once at the lab, their vital signs were monitored. The first test was a modified Bruce Protocol on a treadmill which began at 1,7 mph with a 7% incline. The speed and incline were increased every three min until the testing lasted 15 min (or five stages of the protocol), which meant subjects were at their target HR. They did the protocol four times with a different odor condition each time: no odor, jasmine, peppermint, and dimethyl sulfide. After the treadmill session, they completed the NASA Task Load Index and Profile of Mood States again. They also had their vital signs retaken. Every testing session was separated by 48h. | A treadmill was used to measure exercise. | Peppermint odor: better performance and vigor; lower effort, fatigue, frustration, physical and temporal workload. Dimethyl sulfide odor: higher physical workload. No odor effect on mental workload/oxygen saturation /pulse/ blood pressure/ mean arterial pressure. No difference between gender. |
| Raudenbush et al., 2001 [11] | United States | Experimental | 40 | 20 | 20 men and 20 women from different intercollegiate sports that required a lot of running. | The testing protocol consisted of four tasks: hand grip (dominant hand), 400-meter dash, push-ups till exhaustion, and 20 basketball free-throw shots. This protocol was performed twice in a randomized order, one time with peppermint oil and one time without odor. There was a minimum of two days of rest between each condition. Each odor condition had an adhesive strip under the patient's nose. | Dynamometer, time | Differences between the two conditions for push-ups and 400-meter runs. The hand grip test was almost significant. No significant differences in the basketball free-throw shots. |
| Pournemati et al., 2009 [12] | Iran | Experimental | 36 | 21.17 | Elite women soccer players | Same conditions for all subjects: Participants were asked to identify three different odors. If they had no olfactory problems, they did a maximal treadmill test followed by a 15 min rest and a body composition test. Then, participants were divided randomly into three groups (peppermint, peppermint and ethanol, and control). Each participant received a cotton pad with the odor corresponding to their group, and then it was placed under their nose. Right after, participants performed the Bruce protocol on the treadmill. | During exercise tests, data were collected with a gas analyzer and a polar HR monitor. | No significant results for the maximal HR and the other variables between groups. |
| Nagai et al., 2000 [13] | Japan | Experimental | 26 | 18-24 | Students from different schools | Participants were divided into the odors group and the control group. Participants in the odors group had to choose a pleasant odor of their choice and inhale it one min before the exercise tests. Participants from both groups rested for 20 min while sitting, and then, two min before the test, data were collected. Then, they did the rhythmic handgrip test, which consisted of maintaining a 200 mmHg pressure till exhaustion by making multiple contractions. When participants stopped, data were collected. Participants also had to complete a static handgrip test which consisted of maintaining the power of 30-40% of their MVC. | Blood pressure monitor, polygraph system | Lower results for the odor group for the diastolic blood pressure in the rhythmic handgrip test. No significant differences between the two groups for the other data and the static handgrip test. |
| Kwon et al., 2020 [14] | Korea | Experimental | 20 | 22.5 | University students participating in regular aerobic exercise without olfactory disabilities | Same conditions for all subjects: Each participant completed a maximal exercise test on a treadmill. Participants had to complete two experimental visits separated by seven days. The first visit consisted of running 30 min on the treadmill without odor stimulation. The second visit had the same treadmill protocol, but participants received an odor stimulation. Feelings and physical arousal were assessed before, during and after exercising. | Questionnaires and face-to-face interviews. Data were collected before, every five min during exercise and ten min after. | Improvement in the feelings of participants when the odor was presented. No significant changes in physical arousal between the two experimental visits. |
| Kim et al., 2018 [15] | Korea | Experimental | 7 | 24.8 | Athletes with a patellofemoral pain syndrome diagnostic | Participants were divided into two groups (control and odor group). Both groups performed the PA program that contains isotonic exercises. For the odor group, participants applied an oil mixture (lavender, rosemary, and peppermint) added to jojoba oil on their wrists. Between each set, participants inhaled the odor. Data were collected before and after the application of the odor. | RPE and blood fatigue markers (ammonia, CPK, LDH) | After the isotonic exercises, lower RPE for the odor group than for the control group. Both groups had an increase in blood ammonia and CPK. For blood LDH, the control group had an increase, while the odor group did not have a significant increase. |
| Kamimura et al., 2009 [16] | Japan | Experimental | 9 | 19-24 | Healthy men college students from Japan | Same conditions for all subjects: No drinking was allowed the day before the experimentation. The session was divided into three parts. First, participants had to rest for six min. Then, the second part was a moderate incremental physical exercise on the bike for 12 min. The last period consisted of resting for 12 min and completing the odor tests. | Questionnaires | Respiratory variables (RF, VT, VE, VO<sub>2</sub>, VCO<sub>2</sub>, oxygen concentration) | Odor impression: Lemon was rated more pleasant than isovaleric acid.<br>Normal breathing during the recovery period: For the isovaleric acid odor, RF decreased at 8-12 min, an increase in VT at 6-12 min and an increase in VCO<sub>2</sub> at 6-8 min. Nose-to-mouth breathing during the recovery period: For the lemon odor, there was an increase for RF at 0-2 min, VE at 6-12 min, VO<sub>2</sub> and energy expenditure at 8-10 min. For the isovaleric acid odor, there was an increase in almost all variables at 6-10 min. There was also a decrease in VT at 0-2 min. |
| Jaradat et al., 2016 [27] | Palestine | Non-randomised quasi-experimental study | 20 | M. spicata oil group: 19.1<br>C. sinensis oil group: 19.8 | Male university students from An-Najah National University in physical education faculty. | Same conditions for all subjects for the pre-test: Participants had to run 1500 meters and complete the lung function tests. Post-test: Participants were divided into the M. spicata oil group and the C. sinensis flower oil group. Each group inhaled five min before the exercise test the group's specific odor that was diffused in the room. Then, participants ran for 1500 meters and completed the lung function tests again. A three-day pre- and post-test interval was given to limit the fatigue impact. |  | Spirometer for the lung function tests and timer for the 1500-meter run | Amelioration in lung function for both groups (increase in FEV1 and FVC), but no significant FEV1/FVC ratio results. Amelioration in the running time for both groups in the post-test versus the pre-test. |
| Hirsch et al., 2007 [17] | United States | Experimental | 20 | 16.1 (14-17) | Women | Participants had to play two rounds of bowling. In each round, participants had to wear a surgical mask that smelled either jasmine or nothing. The order of which mask for which round was random. They also had to rate the sensation of the mask on an analogue scale. | Analogue scale | Bowling score | Amelioration when inhaling the jasmine odor. No differences in the sensation rating. |

|  |  |  |  |  |  |  |  |  |
| --- | --- | --- | --- | --- | --- | --- | --- | --- |
| Esfangreh et al., 2011<br>[18] | Iran | Experimental | 20 | 22-30 | Athlete students that participated in the Championship plays of Iran students 2004-2005. | Same conditions for all subjects: Reaction time test, back muscles strength test, sit-up test and Bruce test were performed before being divided into two groups (control and experimental). Experimental group: Inhalation of the peppermint odor with white alcohol (pumped in the experiment room). Participants did the same tests as pre-inhalation. Control group: Inhalation of white alcohol only. Participants did the same tests as pre-inhalation. | Oxygen consumption, repetitions | No significant relations between pretest and post-test for the control group and the experimental group for the VO<sub>2</sub>max, abdominal muscle strength, and back muscle strength. Difference between pre-test and post-test reaction time for the control group and the experimental group. Effect of peppermint on the VO<sub>2</sub>max. |
| De Pauw et al., 2017<br>[19] | Belgium | Experimental | 11 | 22 | Healthy and active males with a low daily intake of caffeine. | Same conditions for all subjects: They came to the lab seven times. The first time, they did a maximal physical test to obtain their VO<sub>2</sub>max and consulted with a doctor. The participants got familiar with the protocol and nasal spray for two other visits. The trial was the same the next three times except for the changed nasal spray solution. The spray was always administered before the Wingate test and every 7,5 min of the time trial test. They did a warm-up and then performed two exercises: a Wingate test and a 30-min time trial performance. The sprays used were caffeine, glucose, or placebo. At the seventh visit, they collected two blood samples 20 sec after administration of the caffeine spray to measure caffeine concentration. Subjects also performed five Stroop tasks during each experimental visit. | A cycle ergometer was used to measure peak power and average power. | Glucose spray increased average power during the time trial test. No change with caffeine spray for peak power or cognitive task. |
| Cruz et al., 2012 [21] | Philippines | Experimental | 10 | 18.9 (17-21) | Female collegiate basketball athletes with no injury and taking no medication. | Same conditions for all subjects: Participants were not allowed to wear perfume/ other substances that might change the odor during the tests. No vigorous PA 24 hours before each experimental test. Each odor (lavender and peppermint) was randomized between every participant. Participants did a warm-up and a four-min active-rest interval each session while answering the pre-subjective vitality questionnaire. Then, participants had an adhesive strip which contained the correct odor for their group under their nose, and they had to complete the post-subjective vitality questionnaire one minute later. Afterwards, participants did the speed and agility test two times with a four-min rest between. | Timer for the speed and agility tests (same evaluator through the three sessions) | Effect for the lavender group, which was faster and more agile than the peppermint group. No significant differences between the lavender group and the control group in speed, agility, and subjective vitality. The peppermint group felt more vigorous and energetic than the two other groups. |
| Cruz et al., 2011 [20] | Philippines | Experimental | 104 | 18.875 (16-28) | Right-handed, healthy university students with no experience in dart throwing. | Same conditions for all subjects: randomization of all participants in three different groups (lavender, peppermint, control). Each participant received a cotton bud with the odor associated with their group. Participants had to split the bud and put each piece under the nostrils. They inhaled the odor for two min and then filled the State-Trait Anxiety Inventory before finishing with a dart-throwing test. This test had three practice throws followed by 20 throws. | The target was 173 centimeters over the floor, and 237 centimeters from the participant. To assess the scoring, a Cartesian coordinate system was used. | Difference between groups: the peppermint group was more accurate and constant than the lavender and the control group. Less anxiety in the lavender group than in the peppermint and the control group. Higher anxiety level in the control group. No differences between the lavender and the control group for accuracy and consistency. |
| Campbell et al., 2021 [22] | UK | Experimental | 14 | 20 | Trained (non-resistance) male without neuromuscular/musculoskeletal disorders (past and present) | Same conditions for all subjects: no exercise, alcohol, and caffeine 24 hours before the visit; similar diet for the past 48 hours; fasting one hour before the visit. Three visits (control, experimental and placebo/sham) with a rest of seven days between each. The order of the visits was randomized.<br>Experimental visit (AI): Firstly, participants rested for 20 min, and then there was the ammonia inhalation. Before and after each inhalation, HR and alertness were measured. Ammonia inhalation was repeated before each repetition. Participants needed to complete a knee extension, a handgrip maximal voluntary isometric contraction with the dominant limbs, and a bilateral countermovement jump. Each exercise needed to be completed for three repetitions. A 20-min rest separated each exercise, and a 5-min rest separated each repetition. At the end of the experimentation, participants needed to rate themselves on their perceived performance during each exercise. Sham visit: same protocol as the experimental one, but with inhalation of water. Control visit (CON): same protocol as the experimental one, but without odor inhalation. | Isokinetic dynamometer (Peak power/Peak Strength), Polar monitor (HR), Visual Analogue Scale (Alertness) | HR, alertness and perception of performance were higher after ammonia inhalation compared to the sham condition. Positive correlations between alertness and perceived performance after ammonia inhalation. No change in the peak power and reaction time. A significant effect for the rate of force development between 20 and 200ms for the knee extension and handgrip exercise. |
| Becker et al., 1995 [23] | United States | Experimental | 20 | 60-101 | 10 men and 10 women (members of community-wide senior service facility) | Same conditions for all subjects: questions about pre-existing conditions that might exclude them from the research. Nine tests: They needed to walk 90 seconds on an indoor course (61 meters) in different randomized conditions (music and/or odors). They needed to go as far as possible during the time interval. The odor conditions were no odor, chamomile scent, or peppermint scent. Each odor condition was matched with all music conditions: frenetic music, mellow music or white noise. | Measure of walking distance in different conditions on a 61 meters indoor course. | Effect for the music (lower result for the mellow music than the white noise and the frenetic music), but none for the odors. |
| Basevitch et al., 2011<br>[24] | United States | Experimental | 76 | 24.1 | Students from 2 South-Eastern universities | Same conditions for all subjects: Participants had to complete a questionnaire about their global health (GHLQ questionnaire), and they had to complete a hand grip task to measure their MVC. Then, participants were randomly divided into four groups: control, placebo, peppermint, and lavender. All the participants, except for the control group, placed an adhesive strip with the corresponding odors under their noses. Right after, participants had to do the hand grip test again, but at their 30% maximum, until exhaustion. Every 30 sec during the test, they answered questionnaires about the smell, RPE and attention. | A handgrip dynamometer has been used to measure the MVC. | Effects for the pleasantness of the smell in which placebo condition was less pleasant than odors groups. Also, the lavender condition diverted more attention than the peppermint and placebo condition. There were no significant differences between groups for the handgrip duration and the RPE. |
| Wooller et al., 2015<br>[25] | UK | Experimental | 29 | 25.6 | Healthy subjects | Same conditions for all subjects: Participants did a cycling test to find their peak power at the first visit. Then, at the second one, they were randomly put in one of the three sensory-occlusion conditions (visual, auditory and olfactory). All participants did a warmup for five min at 20% of their peak power. When they retrieved their normal HR, they did five min of cycling at 40% of their peak power while watching a video of cycling in the wood with forest sounds and smells. Then, while the participants rested until reaching their resting HR, one sense was obstructed depending on the condition they had been assigned to (visual, auditory, or olfactory occlusion). Then they got back to cycling for five min. They repeated three times the protocol of switching between the sensory-occlusion condition (always the same) and the full sensory condition. | A cycling ergometer has been used to measure RPE, HR and total mood disturbance. | A diminution of mood and higher RPE and HR with sensory occlusion. Even more when sound was occluded. |

CPK: creatine phosphokinase; FEV1: forced expiratory volume in the first second; FVC: forced vital capacity; HR: heart rate; LDH: lactate dehydrogenase; MSIS-29: 29 items Multiple Sclerosis Impact Scale; MVC: maximal voluntary contraction; PA: physical activity; RF: respiratory rates; RM: repetition maximum; RPE: rate perceived exertion; VCO<sub>2</sub>: carbon dioxide uptake; VE: ventilatory volume; VO<sub>2</sub>: oxygen uptake; VO<sub>2</sub>max: maximal oxygen consumption; VT: tidal volume;

**Table 2.**
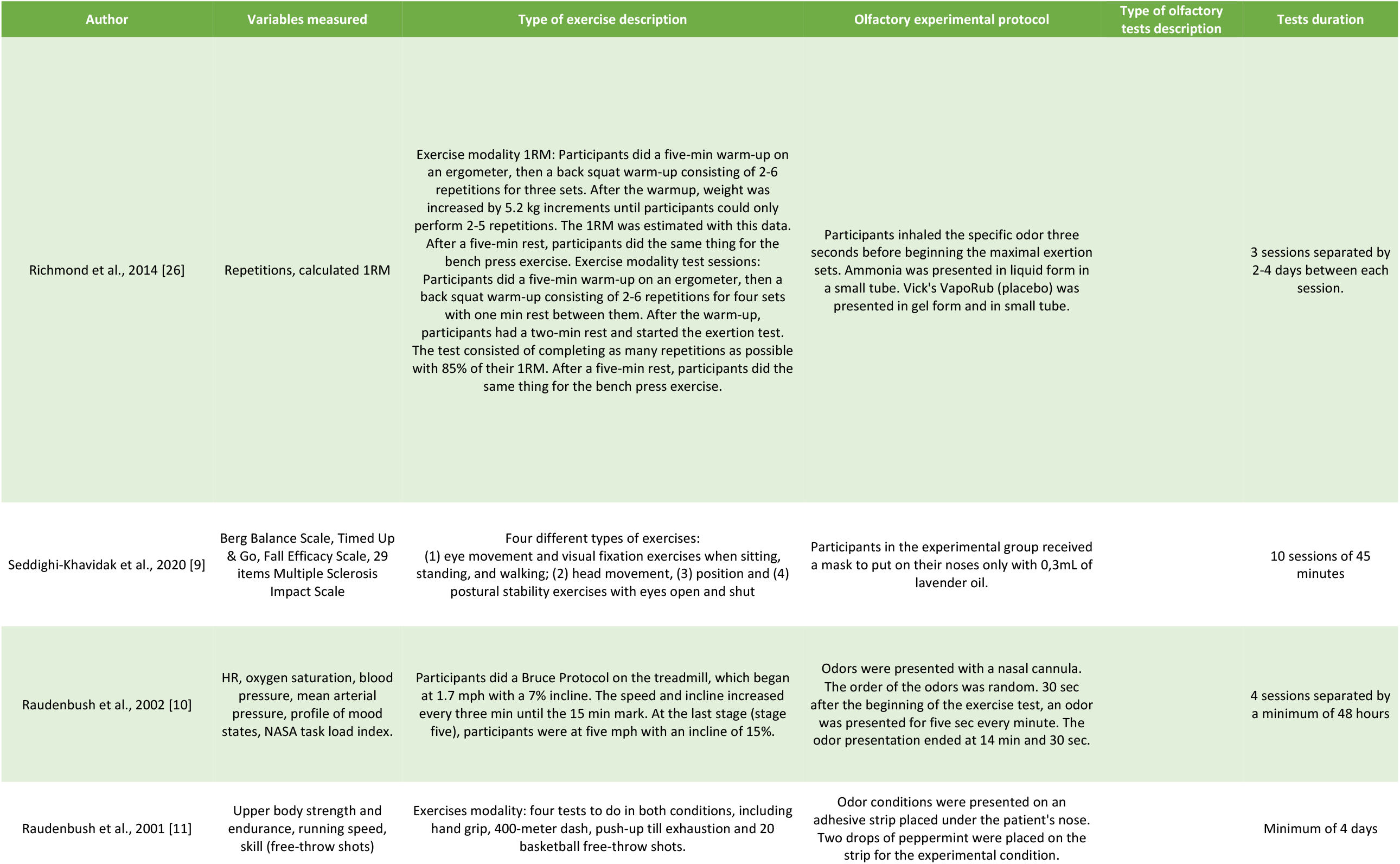
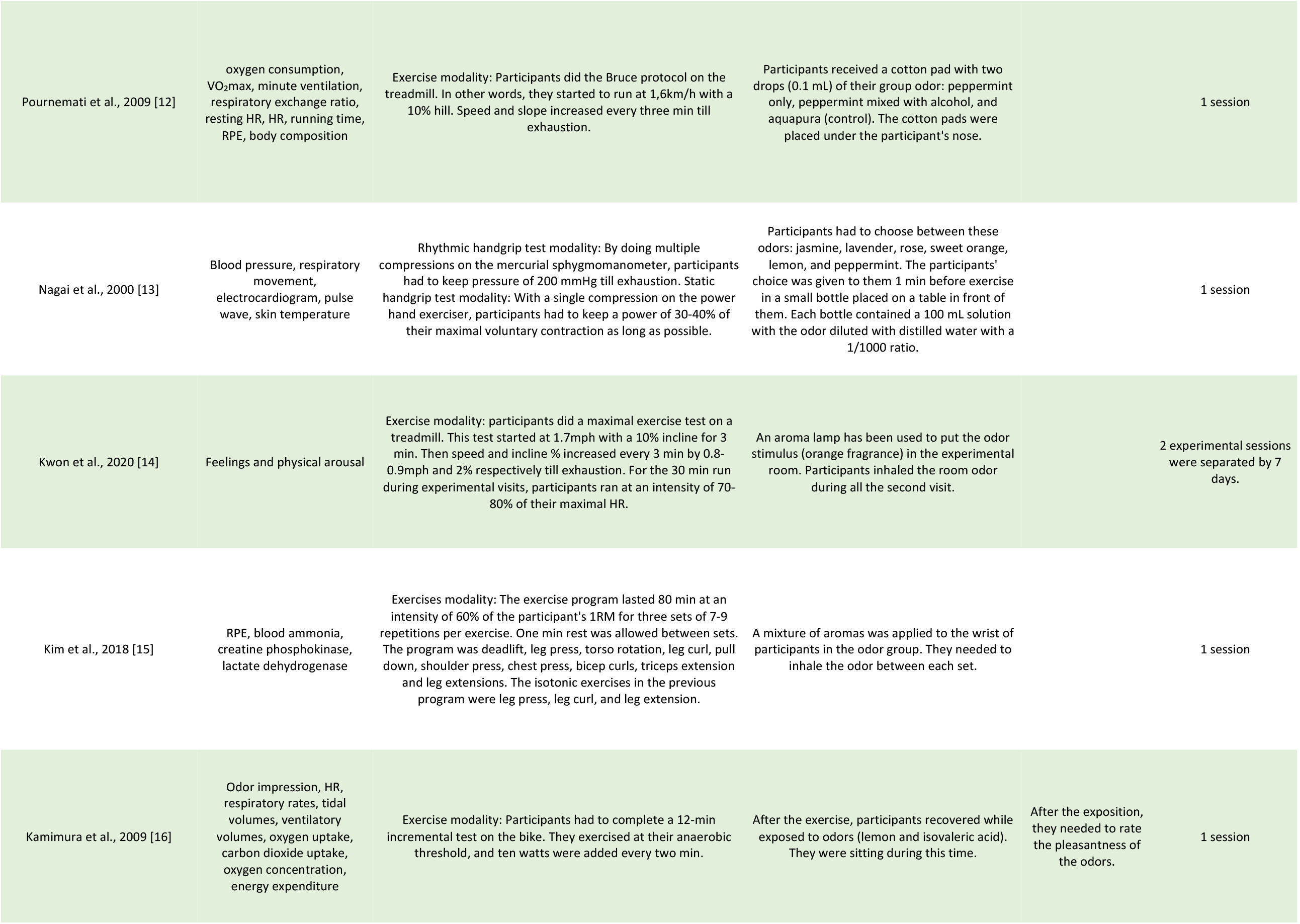
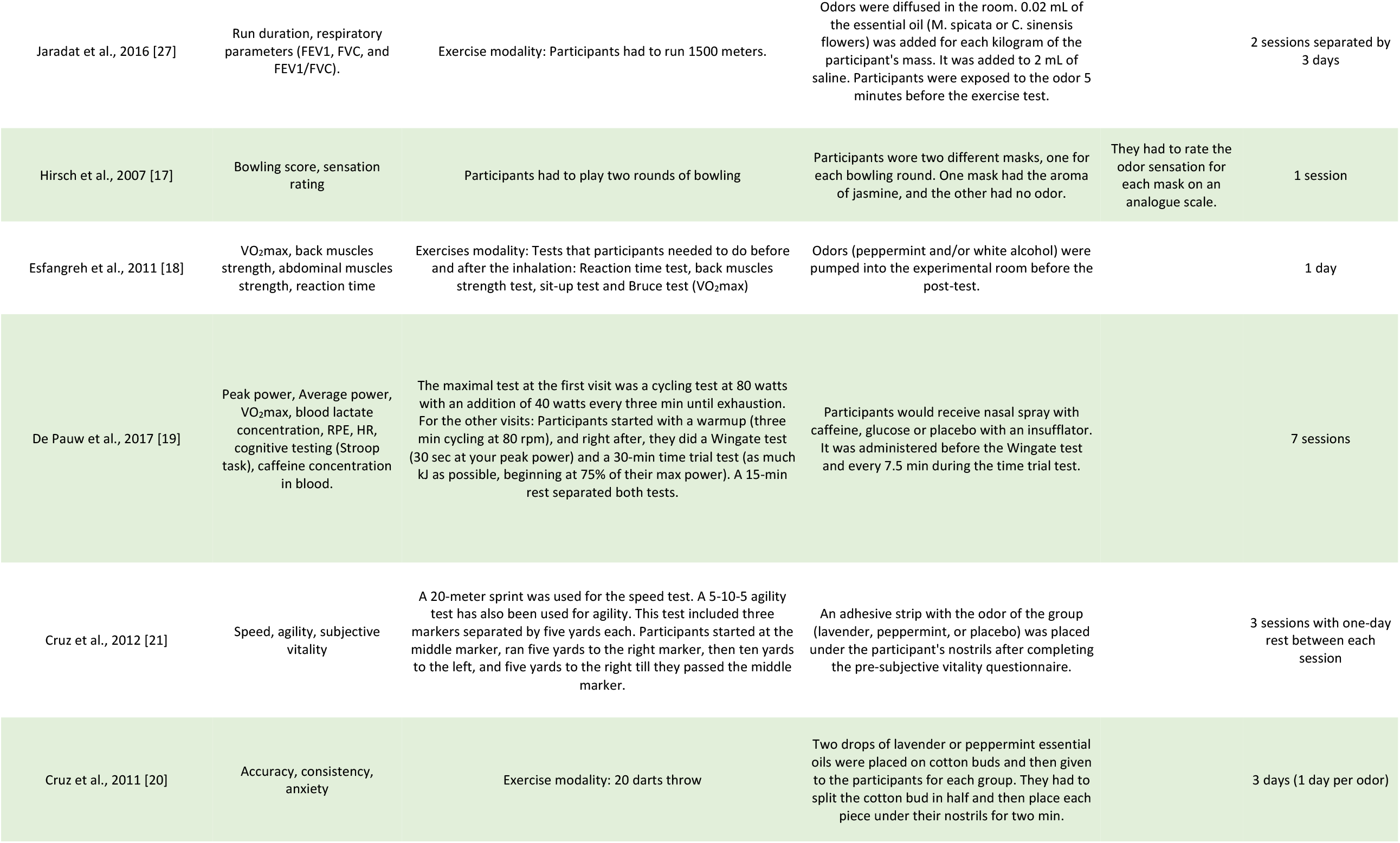
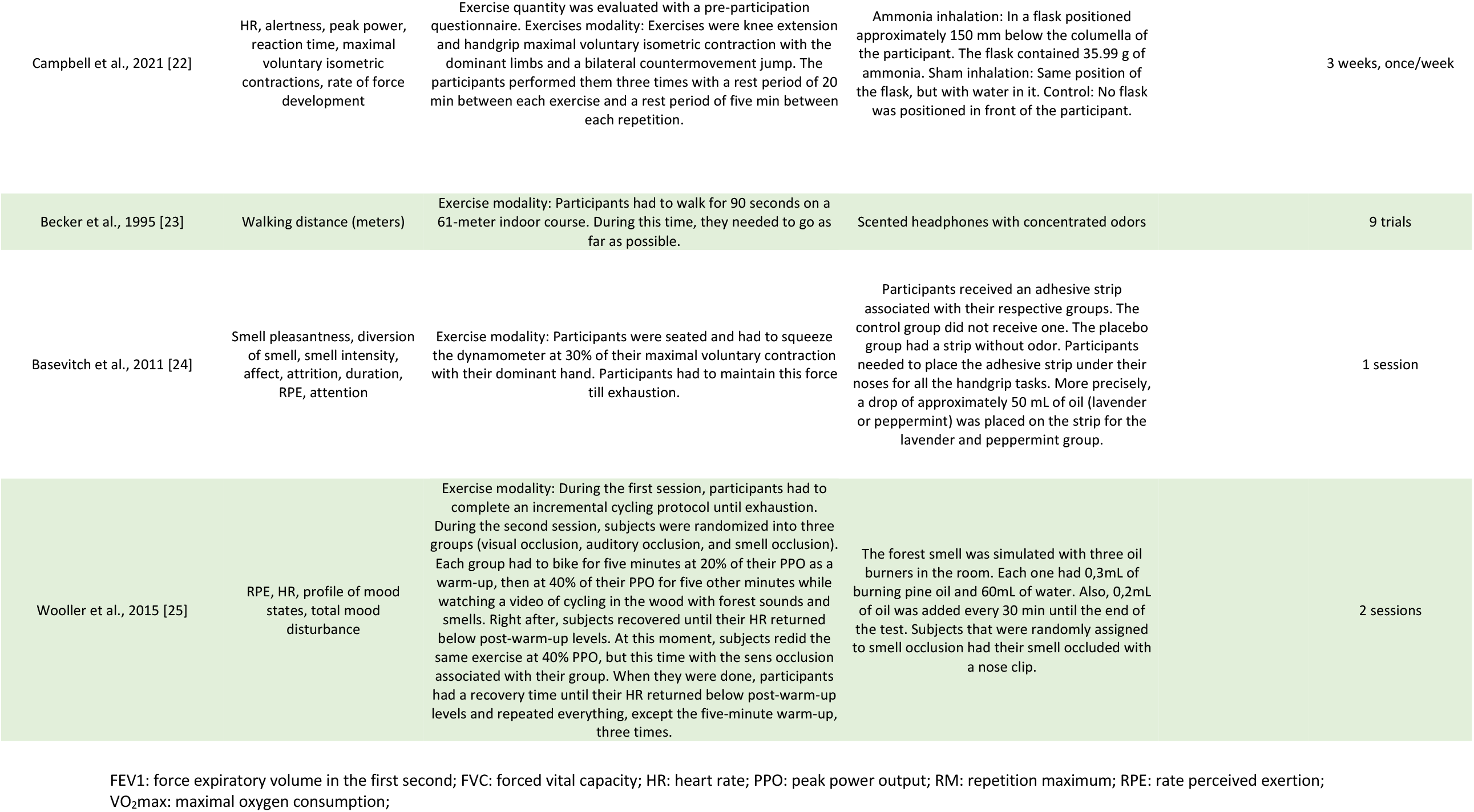
Protocol description of studies included

### 2.4 Risks of Bias and Quality Assessment

Risks of bias and quality assessment were analyzed using the QUADAS-2 tool [8]. This tool consists of four key domains, which cover 1) patient selection, 2) index tests, 3) reference standards, and 4) flow of patients through the study and timing of the index test(s) and reference standard (“flow and timing”) [8]. Each domain was assessed in terms of their risk of bias, and the first three domains were also assessed in terms of concerns regarding applicability. The risk of bias was assessed by one author (ACG), and a consensus was reached through discussion with two authors (MC and AM).

## 3 Results

### 3.1 Study Selection

Two thousand eight hundred seventy-seven studies were screened based on their titles, and 47 were kept for full-text studies assessment. From those studies, 19 studies were analyzed and extracted. As presented in Table 1, those studies had between 7 and 104 participants, for a total of n = 421 participants. Out of these 19 studies, 18 had an experimental design [9-26], while one study had a quasi-experimental nonrandomized design [27]. All 19 studies assessed the effect of certain odors on PA characteristics like strength, cardiovascular capacity, precision, and balance.

### 3.2 Study and Olfactory Interventions Characteristics

As presented in Table 1, studies took place in Belgium [19], Iran [9, 12, 18], Japan [13, 16], Korea [14, 15], Palestine [27], Philippines [20, 21], the United Kingdom [22, 25], and the United States of America [10, 11, 17, 23, 24, 26]. Most of those studies were conducted on young athletes [10-22, 24-27] and the rest on adults [9, 23]. Each study had an intervention time ranging from 1 [12, 13, 15-18, 24] to 10 [9] sessions. Various methods were used to assess olfactory responses. Five studies placed the odor right under the nose with an adhesive strip [11, 12, 20, 21, 24] while other used small tubes [13, 22, 26], nasal spray [19], scented headphones [23], nasal cannula [10], odor on the wrist [15], odor in a mask [9, 16, 17], or odor diffused in the testing room [14, 18, 25, 27]. The period in which the participant inhaled the odor also varied between three seconds [26] to five minutes [27] before the PA. Odors that were presented more often in those 19 studies were peppermint [10-12, 18, 20, 21, 23, 24, 27], lavender [9, 20, 21, 24], citrus [14, 16, 27], and ammonia [22, 26]. There were also jasmine [17], essential oil mixture [15], and forest smell [25]. Caffeine and glucose were used for the nasal spray study [19].

### 3.3 Physical Activity Protocols Characteristics and Tests

As presented in Tables 1 and 2, PA included cardiovascular exercise on a treadmill [10, 12, 14, 18] or a track [11, 21, 23, 27], or cycling on an ergometer [16, 19, 25]; resistance training (e.g., squats, deadlift, leg press, push-up, handgrip) [11, 13, 15, 18, 22, 24, 26]; precision task (e.g., dart throw, basketball free throw) [11, 17, 20]; and/or balance tasks [9]. The effect of each odor was assessed with the changes of some PA variables such as the number of repetitions, rated perceived exertion, effort perception evaluated with different scales, cardiac output, exercise duration, skills ability, and blood analysis. In short, these studies assessed strength, cardiovascular, precision, and balance variables.

### 3.4 Effect of Odors on PA

#### Strength tasks

Out of the 19 studies, seven explored the effect of the odors on strength variables during a resistance training task [11, 13, 15, 18, 22, 24, 26]. Two showed no significant effect of ammonia and peppermint odor on the number of repetitions, the 1RM, the abdominal and back muscles strength [18, 26]. The other five studies reported significant effects: peppermint increased the number of push-ups (+2.2) [11], lavender diverted more the attention (i.e., participants were less focused on the specific task) [24], and ammonia generated a higher heart rate, alertness, and performance perception (i.e., how the participant thought he performed) [22]. The study using oil mixture [15] also showed a lower rated perceived exertion during an isotonic exercise (e.g., participants rated 2.1 lower on the RPE scale with odor inhalation during the biceps curl exercise). Finally, the study in which the participant selected by himself a pleasant odor before testing [13] demonstrated a lower diastolic blood pressure (6.7 mmHg lower) during a rhythmic handgrip task.

#### Cardiovascular tasks

Eleven studies assessed cardiovascular variables such as heart rate and blood pressure [10-12, 14, 16, 18, 19, 21, 23, 25, 27], and two of them had no significant results using peppermint odor [12, 23]. However, peppermint seems to increase performance (e.g., 400m run faster by 2.94 sec) [10, 11, 27], VO2max [18], lung function [27], vigorous feeling [21] and seemed to decrease fatigue [10] and physical workload [10] in five studies. Other odors, such as lemon and orange, improved the well-being feeling of the participant during the PA [14, 16, 27]. The study using nasal spray also showed a significant increase of 2.4% in the average power during a time trial task while using the glucose spray [19].

#### Precision tasks

Two studies explored the effect of odors on precision tasks. All but one showed a significant effect [17, 20], and one study produced no significant results [11]. The main results from those studies were an increased score of 26.5% during a bowling game when inhaling jasmine odor [17] and better accuracy and constancy when using peppermint odor during dart throws. Lavender decreased participants’ anxiety levels [20].

#### Balance tasks

Only one study assessed the effect of odors on a postural balance task [9]. When using lavender oil, participants had better results on the Fall Efficacy Scale and Timed Up and Go tests.

### 3.5 Risk of Bias Assessment

Although most of the studies included in this systematic review do not have any ‘‘risk of bias’’ nor ‘‘applicability concerns’’, some cases were deemed unclear or at high risk. Results are summarised in Table 3. One study by Esfangreh, Azarbaijani [18] raised too much uncertainty and concerns about its applicability and especially about its risk of bias. It thus made it ineligible to be included in the present systematic review. Even though the study by Hirsch, Ye [17] also presented some unclear risks of bias or applicability concerns, its methodology and presentation made it eligible for analysis in the present systematic review. Concerning patient selection, out of the remaining 17 studies, three had an unclear risk of bias [10, 11, 16]. Concerning the index test, none of the 17 remaining studies had an unclear or high risk of bias. Concerning reference standards, three studies had an unclear risk of bias [19, 21, 23]. Concerning flow and timing, one study had an unclear risk of bias [16]. For the applicability concerns, four studies had an unclear risk for patient selection [10-12, 16]. Concerning the index test, one study had an unclear risk [23]. Concerning reference standards, two studies had an unclear risk [21, 23]. Overall, only two studies presented a questionable risk of bias assessment and quality assessment results [17, 18], and one was deemed unfit for further analysis [18].

**Table 3.** Risk of Bias Assessment [8]

| Author, year | Risk of Bias |  |  |  | Applicability Concerns |  |  |
| --- | --- | --- | --- | --- | --- | --- | --- |
|  | Patient Selection | Index Test | Reference Standard | Flow and Timing | Patient Selection | Index Test | Reference Standard |
| Richmond et al., 2014 [26] | (+) | (+) | (+) | (+) | (+) | (+) | (+) |
| Seddighi-Khavidak et al., 2020 [9] | (+) | (+) | (+) | (+) | (+) | (+) | (+) |
| Raudenbush et al., 2002 [10] | (?) | (+) | (+) | (+) | (?) | (+) | (+) |
| Raudenbush et al., 2001 [11] | (?) | (+) | (+) | (+) | (?) | (+) | (+) |
| Pournemati et al., 2009 [12] | (+) | (+) | (+) | (+) | (?) | (+) | (+) |
| Nagai et al., 2000 [13] | (+) | (+) | (+) | (+) | (+) | (+) | (+) |
| Kwon et al., 2020 [14] | (+) | (+) | (+) | (+) | (+) | (+) | (+) |
| Kim et al., 2018 [15] | (+) | (+) | (+) | (+) | (+) | (+) | (+) |
| Kamimura et al., 2009 [16] | (?) | (+) | (+) | (?) | (?) | (+) | (+) |
| Jaradat et al., 2016 [27] | (+) | (+) | (+) | (+) | (+) | (+) | (+) |
| Hirsch et al., 2007 [17] | (+) | (?) | (?) | (?) | (+) | (?) | (?) |
| Esfangreh et al., 2011 [18] | (?) | (-) | (?) | (?) | (?) | (?) | (?) |
| De Pauw et al., 2017 [19] | (+) | (+) | (?) | (+) | (+) | (+) | (+) |
| Cruz et al., 2012 [21] | (+) | (+) | (?) | (+) | (+) | (+) | (?) |
| Cruz et al., 2011 [20] | (+) | (+) | (+) | (+) | (+) | (+) | (+) |
| Campbell et al., 2021 [22] | (+) | (+) | (+) | (+) | (+) | (+) | (+) |
| Becker et al., 1995 [23] | (+) | (+) | (?) | (+) | (+) | (?) | (?) |
| Basevitch et al., 2011 [24] | (+) | (+) | (+) | (+) | (+) | (+) | (+) |
| Wooller et al., 2016 [25] | (+) | (+) | (+) | (+) | (+) | (+) | (+) |

(+)—Low Risk; (-)—High Risk; (?)—Unclear Risk.

## 4 Discussion

The objective of this systematic review was to document the effect of odors on various types of PA. Out of the 19 studies selected, 18 were retained after the risk of bias assessment, in which Esfangreh’s study was removed [18]. The odors from the most to the least used for the tests were peppermint [10-12, 20, 21, 23, 24, 27], lavender [9, 20, 21, 24], citrus [14, 16, 27], ammonia [22, 26], forest smell [25], jasmine [17], an oil mixture [15], a glucose spray [19] and any participant’s choice odor [13]. With odor interventions, five studies out of six showed significant improvement in some parameters (e.g., 1RM, number of repetitions, abdominal and back muscles strength, participants’ attention, cardiac output) during a strength task [11, 13, 15, 22, 24]; eight out of ten for a cardiovascular task [10, 11, 14, 16, 19, 21, 25, 27]; two studies out of three also demonstrated an increase in precision [17, 20]; and an increase in postural balance was also tested and reported in one study [9]. Overall, this systematic review demonstrated that some odors might improve PA performance depending on the odors’ exposition.

### 4.1 Strength Tasks

Six studies that explored the effect of odors on strength tasks were included in this review, and four of them used a stimulating odor, including peppermint and ammonia [11, 22, 24, 26]. One study had no significant results with ammonia while trying to test if there was a strength increase by measuring the 1RM and the number of repetitions at 85% of the 1RM [26]. The other studies showed a significant increase in the number of push-ups (+2.2) (peppermint) [11], heart rate (+11.5%) (ammonia) [22], alertness (ammonia) [22], performance perception (ammonia) [22], and pleasantness (peppermint) [24]. These significant results are in line with the current literature. Some studies explained that menthol, an organic compound from peppermint, would decrease the discomfort during breathing and may be caused by its stimulation of the sensory and palatine nerves [28-30]. In contrast, even though ammonia is also a stimulant odor, its effect comes from another pathway. Indeed, for ammonia odor, some studies have postulated that it would be the irritation of the lungs and the nose in response to the stimulating odor inhalation that would triggers an inhalation reflex, causing rapid inhalation. It would then results in better and faster work from the breathing muscles, causing an increase in alertness [31, 32]. This process might explain the results obtained in both studies, while the non-significant one [26] evaluated the strength and the significant one [22] the participants’ feelings during the strength task. Since the PA were strength tasks, the respiratory system was less solicited than for other types of PA. Therefore, it may explain why the participants’ feelings are better, but not the strength itself.

In three studies, different odors were used [13, 15, 24]. More precisely, one study let their participants decide their pleasant odor to inhale, resulting in a decrease in their diastolic blood pressure during a rhythmic handgrip task (e.g., 6.7 mmHg lower) [13]. Another study used an oil mixture containing lavender, rosemary, and peppermint. It appeared that participants in the odor intervention group had lower rate of perceived exertion on the Borg Scale from 6 to 20 during the exercises (e.g., 2.1 lower during biceps curl exercise and 1.4 lower during leg curl exercise) [15]. While it may be possible to hypothesize that the oil mixture is a pleasant odor with the use of rosemary, multiple studies have suggested that pleasant and unpleasant odors might activate different pathways in the peripheral and central nervous system [33, 34]. It may also cause a change in the heart rate, where pleasant odors would decrease its frequency and therefore decrease blood pressure [33, 35]. Finally, the last study showed that lavender inhalation have decreased participants’ attention during a handgrip task, which is the first significant negative outcome during a strength task [24]. However, those results are not in agreement with the literature on this odor. A systematic review has suggested that attention should increase following the inhalation of lavender [36]. The authors of this review supported their statement with the linalool, one of the main components of the lavender essential oil, and its effect on the GABAergic pathway [36]. Knowing that participants in the study with decreased attention had the same adhesive strip under their nose for the hole experimentation, there might have been a familiarization effect or a diminished linalool concentration at the end of the task. These results suggest that each of the odors described has the potential to improve performance during strength exercise. However, it is important to note that each odor affects specific parameters of strength tasks and participants’ feelings.

### 4.2 Cardiovascular Tasks

Cardiovascular tasks were explored in ten studies included in this review, six of them used the peppermint odor [10-12, 21, 23, 27]. Out of these six studies using peppermint, four showed a better overall performance with faster running time (e.g., 2.94 sec faster for 400m run) [10, 11, 27], two better participants’ feelings evaluated with different scales (e.g., effort, fatigue, frustration) [10, 21], and one improved lung function evaluated with a spirometer [27]. Similarly, as described earlier in this review, peppermint and, more precisely, menthol might decrease breathing discomfort, which explain such improvements [28-30]. Two other studies produced no significant results [12, 23]. One study with nonsignificant changes in the maximal heart rate with the peppermint odor [12] is also coherent with the ambiguous literature that reported no change in heart rate with nasal administration of this substance [37]. The last study without significant results [23] measuring walking distance after inhaling peppermint might be explained by the intensity of the exercise that was submaximal (walking: 1.5-3.8 METS) in comparison to other studies which had more intense exercises during their experimentation (running: 3.5-23 METS; bicycling: 3.5-16 METS) [38].

Three studies tested citrus odor during aerobic exercise, and they all had significant results [14, 16, 27]. Indeed, participants had a significant increase in feeling (i.e., participants’ mood), which was evaluated with different scales, lung function, which was evaluated with a spirometer, and performance (e.g., 53.20 sec faster for a 1500m run). These results are supported by the literature in which this odor is characterized by its stimulant effect and ability to enhance mood and mental, and physical performances [39, 40]. Some studies have also pointed out one crucial benefit of using this odor which is the lesser number of adverse effects compared to other odors such as unpleasant odors [39, 41].

The remaining three studies used lavender [21], forest smell [25] and a glucose spray [19]. In the first one, participants became faster and more agile after inhaling lavender while performing twice an agility test [21]. Those surprising results might be caused by increased attention as described by Malloggi, Menicucci [36]. It might also be related to the hypotensive effect of this odor that would put the participants in a better mood and a better physiological situation (i.e. lower blood pressure) [42]. It might therefore be suggested that a decreased anxiety during the task enhances its outcome The second study using the forest smell to mimick an outdoor practice of PA has shown that the occlusion of this smell may decrease the participants’ mood and increase the rate of perceived exertion and heart rate [25]. On the one hand, it might be possible that the forest odor was pleasant and would have a similar effect as described earlier for that type of odor although, there is very little literature on this odor. On the other hand, it might also be possible to analyze this result by emphasizing on the occlusion of the sense of smell and the unpleasantness of this situation. Indeed, Johnson [3] has stated that the inhalation of odor can affect self-rating mood measurements positively or negatively depending on the pleasantness of the odor. Another author showed a negative correlation between heart rate and odor pleasantness. [33] These results for unpleasant odor are in line with the research included in this review, but more studies are still needed to better understand the underlying mechanisms. The third study used a glucose nasal spray which increased average power (+2.4%) on an ergometer during a time trial [19]. This effect seems more related to the glucose molecule than the odor itself. Knowing that high-intensity PA can decrease whole-body glucose concentrations in humans [43], better regulation of this substance seems possible with intranasal administration. The study of Pontiroli, Alberetto [44] supports this fact by exposing an increased blood glucose concentration after the nasal administration.

These results suggest that every odor has its advantages and its mechanism. However, the positive effect might be more related to the participants’ mood rather than directly affecting their physical abilities.

### 4.3 Precision Tasks

Three of the 19 studies explored the effect of peppermint, lavender, or jasmine odor on the performance of a precision task [11, 17, 20]. Two of them had significant results [17, 20]. Peppermint generated ambiguous results, with different results reported in two studies [11, 20]. One of these studies noted no improvement of a basketball free-throw accuracy test [11], while the other had a significant increase on a dart-throw accuracy test [20]. Some studies have supported that peppermint increases vigilance and visuomotor accuracy [45, 46]. Therefore, it seems possible that peppermint odor has a positive effect on precision tasks. The differences between these two studies might come from participants’ selection. The study with the basketball free-throw accuracy test [11] recruited highly trained university athletes, including basketball athletes, and the other study showing a significant effect of odor on task accuracy recruited non-experienced university students [20].The benefit of peppermint odor may have a reduced effect on experts than on novices for a precision task.

The same study that reported significant results obtained a lower anxiety score when participants inhaled lavender odor than peppermint or no odor [20]. Even though this odor failed to increase performance, changes in the participants’ mood seem an important variable for precision tasks. A study by Nieuwenhuys and Oudejans [47] showed a negative correlation between anxiety levels and performance during an accuracy task. This lower anxiety level in the study using lavender is supported by the study of Sayorwan, Siripornpanich [48], in which lavender had a similar effect on the anxiety level by positively affecting the parasympathetic system activity. They also used an electroencephalogram for a better understanding of the cerebral changes, and they found an increase in the alpha band which can be interpreted as a relaxed state [48].

The third and last study that assessed a precision task used jasmine odor stimulation and found a better bowling score (+26.5%) while using this odor [17]. Like lavender, jasmine odor was rated as a pleasant odor which might explain its effect on decreasing average heart rate [33, 49]. One study also highlighted higher activation of the parasympathetic nervous system following the inhalation of jasmine [50]. Knowing that a similar mechanism happened with lavender odor and significant improvements were associated with it, more studies using jasmine odor should be conducted to confirm its impact on a precision task. These results suggest that odor stimulation significantly affects precision tasks when these odors are pleasant and presented to participants having little experience in the experimental task.

### 4.4 Postural Balance Tasks

The effect of odors on postural balance tasks has very little literature, with only one study included in this review which used lavender odor [9]. This study reported significant increases in all their postural balance tests, such as Berg Balance Scale, Timed Up and Go, and Fall Efficacy Scale (e.g., improvement of the Fall Efficacy Scale by -3.8 for the experimental group instead of +0.4 for the control group, with lower results showing a reduced fall risk) after inhaling lavender. As mentioned earlier, this odor seems to enhance the participants’ mood by modulating the parasympathetic nervous system [48]. It may also increase the participant’s attention and decrease anxiety [36]. One study assessed the relation between postural balance and anxiety, and it appeared that motor control was more difficult for those who were more stressed [51]. Both studies support each other, with better postural balance in a more relaxed mood.

### 4.5 How Can We Produce More Robust Results?

Studies evaluating the effect of different odors on PA have been creative in their administration methods. Indeed, the duration of the exposition, the distance between the odor and the participant, and the odor concentration were not consistent, and this could partly explain the different results reported across the literature. In the future, using different tools such as an olfactometer might be important to improve the consistency of the olfactory intervention. With the ability to control the duration of the exposition, the distance between the odor and the participant, and the odor concentration, the olfactometer seems the key to more standardized experimental protocols to present odors.

### 4.6 Strengths and Limitations

Even though many odors were tested, peppermint and lavender were the most tested. Those two odors were used for each type of PA included in this review (e.g., strength, cardiovascular, and precision tasks). However, balance tasks only tested lavender odor [9]. It was, therefore, impossible to understand the true mechanism of action of each odor. This systematic review also covered numerous physical qualities (e.g., 1RM, heart rate, overall performance) part of PA with the 19 studies included in this question. By separating each physical quality, a better understanding of the effects of different type of odors was possible. To perform better analyses, other variables than task performance were measured in most studies. These variables helped to generate a better understanding of the mechanisms in action after inhaling different odors.

The main limitation of this review is the irregularity in odor concentration and the duration of the odor stimulus. For example, duration of exposition range from 3 sec [26] to the total duration of the exercise [9, 11, 14, 17, 23, 24]. Knowing that some odors, such as peppermint and lavender, have a specific compound that plays a crucial role in the effects described earlier, their concentration should be standardized to assess the magnitude of their effects [28, 36]. Finally, another limitation of this review is the population sample. Most studies recruited healthy and/or athletic university students. As mentioned in a systematic review made by our lab recently, chronic PA change odor perception [52]. Therefore, it is important to have homogeneous groups in term of physical quality. Also, since olfaction ability seems to decrease with ageing [53-58], it could be interesting to test if the effect of odor stimulation on PA may be affected by age. All those inconsistencies in the methods made the possibility of doing a meta-analysis impossible. Also, since this review includes all the studies mentioned in this paragraph, the same strengths and limitations apply to this review.

## Conclusion

In summary, certain types of odors, such as peppermint and lavender, seem to improve some aspects of PA performance, such as cardiac output and precision, by modulating the sensations perceived. Concerns about inactive behavior have surfaced in several global organizations, such as the WHO. Therefore, studies evaluating the impact of environmental stimuli, more precisely the impact of odors on the practice of PA, are important to deepen our understanding of this increasing problem in the world.

## Data Availability

All data produced in the present work are contained in the manuscript

## Acknowledgment

The authors thank Denis Arvisais for his help with the search strategy.

## Funding

Mathieu Cournoyer and Alexandre-Charles Gauthier received scholarships from the Université de Montréal. Fabien Dal Maso is a research scholar from the Fonds de recherche du Québec – Santé (Junior 1). Marie-Eve Mathieu holds a Canada Research Chair (Tier 2).

## Disclosure of interest

The authors declare no conflict of interest.

## Notes

### Competing Interest Statement

The authors have declared no competing interest.

### Funding Statement

Mathieu Cournoyer and Alexandre-Charles Gauthier received scholarships from the University of Montreal. Fabien Dal Maso is a research scholar from the Fonds de recherche du Quebec - Sante (Junior 1). Marie-Eve Mathieu holds a Canada Research Chair (Tier 2).

